# *APC* I1307K and clinical management: insights from UK Biobank association analysis of colorectal and other cancer risks in Ashkenazi and non-Ashkenazi whites

**DOI:** 10.1101/2025.04.03.25325166

**Authors:** Sophie Allen, Charlie F. Rowlands, Andrew Latchford, Clare Turnbull, Laura Valle

## Abstract

*APC* c.3920T>A; p.Ile1307Lys (I1307K), prevalent in individuals of Ashkenazi Jewish (AJ) origin, has been associated with a modestly increased colorectal cancer (CRC) risk. Clinical recommendations for I1307K heterozygotes vary across countries and expert groups, reflecting differences in population frequencies, modest risk estimates, and limited data in non-AJ individuals. We analyzed UK Biobank data comprising 466,315 individuals (8,727 with CRC), using genomic analysis to classify AJ and non-AJ ancestries. I1307K was identified in 7.1% of AJ and 0.08% of non-AJ white participants. No significant association with CRC was observed in AJ (OR: 0.71; 95%CI: 0.17–2.95) or non-AJ white individuals (OR: 1.05; 95%CI: 0.50– 2.22). The previously established OR of 1.7–1.8 for AJ individuals lies within our 95% CI, indicating underpowered results due to limited CRC cases. No significant associations were detected for other cancers. Unbiased, adequately powered CRC case-control studies in non-AJ populations would require cohorts far larger than current resources for feasible analysis. Clinical actionability of I1307K should prioritize risk stratification based on overall CRC risk and ancestry-dependent variant detection rates. However, management strategies need not differ by ancestry once a carrier is identified, as the biological impact of I1307K should be consistent across populations.

## INTRODUCTION

The *APC* (NM_000038.6) c.3920T>A p.Ile1307Lys (I1307K) missense variant has been associated with a moderate/low increased risk of colorectal cancer (CRC) in individuals of Ashkenazi Jewish (AJ) descent, in whom the allele frequency is the highest (MAF: 3.7%). The level of CRC risk associated with this variant in the AJ population (OR: 1.7-1.8) is similar to the observed epidemiological risk of CRC for an individual with one first-degree relative affected with CRC. There is no evidence for increased risk of polyposis, and the age at cancer diagnosis in I1307K AJ heterozygotes is similar to the age at diagnosis in the general population (mean age: 60-70 years).^1^

Although *APC* I1307K is predicted benign (AlphaMissense: 0.141; REVEL: 0.233; CADD: 8.522), it is located in the β-catenin binding domain of APC and affects an (A)3(T)(A)4 sequence, converting it into an extended tract of eight adenosine nucleotides (A8). *In vitro* and *in vivo* studies performed in the late 1990s suggest that this modification causes the slippage of the polymerase during DNA replication, conferring an increased propensity for somatic truncating mutations in that allele.^2,3^ Likewise, a single *in vitro* study showed that *APC* I1307K modestly increases β-catenin-regulated transcription; i.e., results in the moderate activation of Wnt signaling.^4^

In 2023, the International Society for Gastrointestinal Hereditary Tumours (InSiGHT) issued an evidence-based position statement on *APC* I1307K and cancer risk based on meta-analyses of available data,^1^ which included consensus recommendations for the clinical management of carriers of I1307K and identification of research gaps that needed to be addressed. The group recommended that clinical CRC surveillance be restricted to carriers of *APC* I1307K of AJ origin on the basis that the only evidence for association with CRC was in the AJ population. The InSiGHT group highlighted the need for well-designed, geographically matched case-control studies to be able to make a clinically informative statement regarding association of *APC* I1307K for non-AJ individuals. The scarcity of data to assess the risk of extracolonic cancers and the lack of robust methods used to define ancestry in the published studies were also highlighted as research priorities.

Here we present an analysis of the UK Biobank prospective cohort study comprising 466,315 individuals, and a genomic principal component (PC) analysis to identify an AJ ancestry group,^5^ aimed to investigate the association of *APC* I1307K with CRC and other cancer types in AJ and non-AJ individuals. Previous studies that evaluate the risk of cancer in I1307K carriers had relied on self-reporting of AJ ancestry, and/or the country of birth of parents and grandparents when Jewish descent was suspected. Here, we used a PC analysis based on genomic data, which enabled systematic assigning of ancestry equivalently to study participants, both AJ and non-AJ and both CRC cases and those unaffected by CRC.

## MATERIAL AND METHODS

### UK Biobank individuals included in the analysis

Full autosomal sequence data was retrieved from all participants within the UK Biobank (UKB).^6^ Related individuals were excluded based on a kinship relatedness cut-off of 0.0884 (second degree relatives or closer) using PLINK2.^5,7^ 466,315 individuals were retained after filtering.

### Ancestry assessment using PC analysis

Data from UKB individuals was projected onto the 25-principal component space of a reference dataset of 202 Jewish individuals and 1,572 non-Jewish individuals using the bigsnpr package.^5,8^ Following methodology described in Prive et al., the robust geometric mean of the Ashkenazi Jewish (AJ) population in the reference dataset was calculated, and the average distance of each Jewish population to this geometric mean generated.^5^ A distance threshold of 15 was deemed sufficient to assign an individual to the AJ group based on inspection of a distance histogram and to maximise the number of AJ reference individuals while limiting the number of non-AJ Jewish individuals. Due to overlap in distance scores, the threshold encompassed 23/29 “Ashkenazi Jewish”, 2/10 ‘Italian Jewish’, 1/6 ‘Tunisian Jewish’, and 1/22 ‘Sephardi Jewish’ based on self-reported ethnicity for individuals in the reference dataset. Decreasing the threshold to include only self-reported Ashkenazi Jewish individuals (threshold=9.2) would only retain 4/29; likewise, increasing the threshold to include more Ashkenazi Jewish references would increasingly include other ethnic groups. A UK Biobank individual was thus classified as ‘AJ’ if the average distance across PCs was equal to or below the distance threshold of 15, and ‘non-AJ’ if above the threshold.

Non-AJ ethnicities were defined using UKB data-field 21000, collapsed using top-level ethnicity (“Asian”, “Black”, “Chinese”, “Mixed”, and “White”). Where ethnicity for an individual indicated multiple of the top-level ethnicities or an ethnicity of “Other”, an ethnicity of “NA” was instead assigned.

### APC I1307K genotyping and cancer data in UK Biobank

Individuals were considered to be carriers of NM_000038.5: APC c.3920T>A; p.(Ile1307Lys) if a variant was extracted from the UKB population-level pVCF-formatted variant^9^ at the GRCh38 chr5 112839514 position with a T>A change, without overlap with deletions or insertions covering this region. Homozygous and heterozygous carriers of this variant were annotated against participant ID and filtered based on AJ vs non-AJ status.

Each phenotype was defined using a combination of the reported ICD9 and ICD10 codes (UKB data-fields 40013 and 40006) and self-reported diagnoses (field 20001). The selected terms searched to assign each cancer phenotype are described in **Supplementary Table 1**.

### Statistical analysis

Fisher’s exact test of independence and quantification of effect size using an odds ratio (OR) were used to test and measure the difference in carrier frequency between phenotype-positive and phenotype-negative individuals for AJ and non-AJ carriers. The OR and 95% confidence intervals (lower 95% confidence interval = LCI, upper 95% confidence interval = UCI) were calculated as follows:

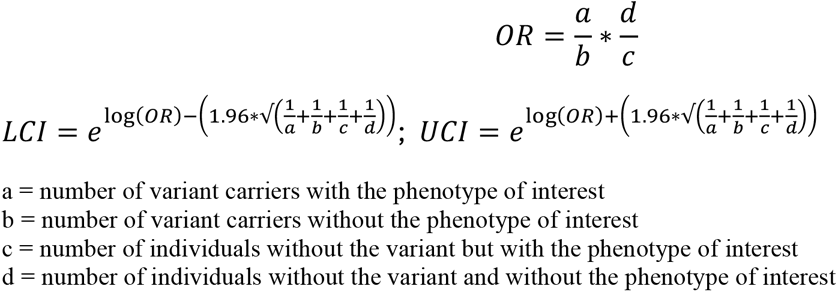

## RESULTS

Using PC analysis, we identified 2,726 individuals as being of AJ ancestry. The frequency of the *APC* I1307K in the UK Biobank AJs was 7.1% (193/2,726), a similar frequency as reported in control AJ populations of published studies.^1^ 436,938 non-AJ white individuals, as assessed by PC ancestry analysis, were identified (**Supplementary Figure 1**). The I1307K frequency in these non-AJ white individuals was 0.077% (336/436,938), a frequency comparable to that reported in gnomAD v.4.1.0 for non-Finnish Europeans (480/589,993; 0.081%). No homozygotes were identified in the non-AJ white populations of either UK Biobank or gnomAD.

We identified 8,727 CRC cases in UK Biobank based on cancer registration and patient self-report, including both previous and prospectively ascertained diagnoses. Within the AJ population, the frequency of *APC* I1307K in those with a CRC diagnosis was 2/39 (5.1%) vs. 191/2,687 (7.1%) in AJ individuals without CRC (OR: 0.71, 95% CI: 0.17-2.95).

When considering non-AJ white individuals, no association of I1307K with CRC risk was identified: 7/8,688 (0.08%) CRC patients vs. 329/428,250 (0.08%) individuals without CRC (OR: 1.05, 95% CI: 0.50-2.22) (**Table 1)**. There was no evidence for association in non-AJ non-white populations of I1307K with CRC or other cancers, either individually or collectively (**Supplementary Table 2**).

**Table 1:**
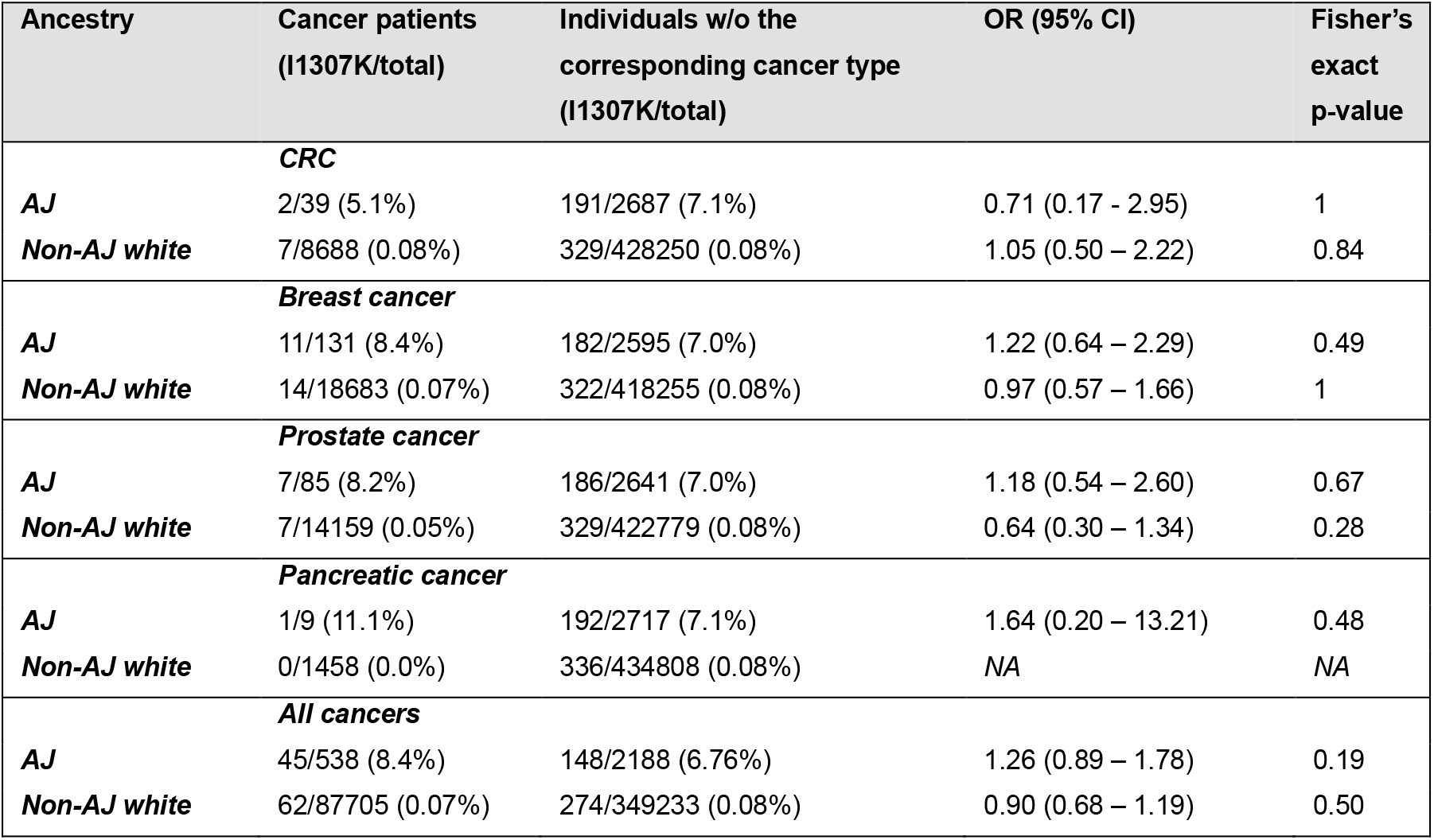
*APC* I1307K frequency in CRC, breast, prostate, pancreatic, and all cancer patients and patients without these phenotypes in the UK Biobank, and associated ORs for AJ and non-AJ white individuals.

## DISCUSSION

This is the first time a population-based prospective cohort study has been used to assess the cancer risks associated with *APC* I1307K. No association with CRC, breast cancer, prostate cancer, or pancreatic cancer was identified for either AJ or non-AJ white individuals (**Table 1**).

The estimate of association of I1307K with CRC in AJ individuals is OR: 0.71 (95% CI: 0.17-2.95), meaning that the previously-established estimate of association of OR: 1.7-1.8 lies within the 95% confidence interval.^1^ The breadth of the confidence interval reflects the comparatively modest numbers of AJ individuals in UK Biobank, particularly the small number of AJ CRC cases, meaning this study is underpowered for analysis of association of CRC in AJ patients compared to the multiple previous case-control studies in AJ individuals. Therefore, this analysis does not inform or alter current understanding of the association of CRC with I1307K in AJ individuals.

The association of I1307K and CRC in non-AJ white individuals has long been a subject of debate, as reflected by the InSiGHT recommendations. In a large case-control study by Forkosh et al.^10^ the frequency of the APC I1307K variant in non-AJ white CRC patients was 64/26563 (0.24%): these were ascertained from clinical genetic testing data from Invitae, a US commercial laboratory, and described as being of self-reported French Canadian or white ethnicity (although the breakdown therein was not delineated). On comparison to non-AJ non-Finnish European controls from gnomAD v.2.1, in whom the frequency of I1307K was 73/58,918 (0.12%), the OR for I1307K and CRC was estimated to be 1.95 (95% CI: 1.39-2.73). The constitution of the gnomAD v.2.1 Non-Finnish European control series is described as: 40% NorthWestern European, 20% Swedish, 9% Southern European, 6% Bulgarian/Estonian and 26% other Non-Finnish European (based on genomically-inferred ancestry). Thus, it is unclear the extent to which population structure (heterogeneity in ancestry between cases and controls) may have influenced the reported association, in particular given the recognised wide variation in frequency for the I1307K variant. Interestingly, this study also reports positive case-control association for the I1307K variant in non-AJ white individuals for melanoma (OR=2.54 (1.57-3.98), female breast cancer (OR=1.73 (1.18-2.65) and prostate cancer (OR=2.42 (1.45-3.94). Notably in the AJ partition of the analysis (using Invitae data for AJ cases of self-reported ancestry and gnomAD v.2.1 for AJ controls), the association with CRC was significant (145/1306 (11.1%) in cases and 342/4918 (6.9%) in controls, OR=1.67 (1.36-2.05)), whilst there was no significant association in any of these other cancers in AJ individuals despite well-powered analyses.

We have presented data from UK Biobank on I1307K in 8,688 non-AJ white CRC cases and 428,250 non-AJ white controls with OR for association between I1307K and CRC being 1.05 (95% CI: 0.50-2.22). The only other study, to our knowledge, of *APC* I1307K in (non-AJ) CRC cases and controls from the same geographical area is a case-control study from Spain comprising 4,072 CRC patients and 2,739 unaffected controls. This also found no evidence of association with *APC* I1307K detected in 7/4,072 (0.17%) CRC patients vs. 4/2,739 (0.15%) controls (OR: 1.18; 95% CI: 0.3-4.0).^1^ It should be noted that no ancestry assessment was performed in this study. Compared to this analysis, our study is more than seven-fold larger overall, having twice the number of cases and 150-fold the number of controls, as well as being a cohort with systematic ascertainment and genotyping of participants agnostic to disease status and ancestry. Nevertheless, the rarity of I1307K in non-AJ white individuals means our analyses still categorically lacks power to demonstrate or confidently refute association of *APC* I1307K with CRC in non-AJ populations. Lack of power was in part driven by the frequency of I1307K in non-AJ white controls in UKBiobank of 0.08% being lower than that reported in the other series (noting technical parameters for calling from WES of I1307K in UKBiobank were of high quality and the frequency of I1307K in the AJ population of 7.1% (193/2726) is wholly consistent with other AJ frequency estimates). Compared to the Spanish series described by Valle et al in which the frequency of I1307K in control population was 0.15%, the lower frequency observed in non-AJ whites from UKBiobank may be a reflection of application of a PCA-based selection of non-AJ whites, and/or due to a remaining-mostly Sephardi-Jewish genetic pool in Spain. Considering that the AJ population in gnomAD v.2.1 was likewise delineated via PCA; the 50% higher frequency of I1307K (0.12%) in the non-AJ non-Finnish Europeans group thus likely reflects heterogeneity in I1307K frequency across non-AJ white populations. This geographic heterogeneity should be considered when reporting I1307K case-control association studies, as previously highlighted by the InSiGHT group.

As mentioned in the introduction, the mechanism of tumorigenesis ascribed to *APC* I1307K lies in the A8 tract, which is prone to mispairing and slippage during DNA replication, thereby creating frameshift changes in that region. The only somatic variant observed in the A8 tract is c.3924_3925insA, which translates into a frameshift change. *APC*, as a classic tumour suppressor, requires two hits to inactivate its tumour suppressor activity, and based on the location of the somatic c.3924_3925insA, loss of the heterozygosity (LOH) of the wildtype allele is expected. Thus, sequencing of colorectal tumour tissue for c.3924_3925insA to identify LOH of the other allele in non-AJ white I1307K variant carriers would also be a potential approach by which to explore aetiologic implication.

However, in AJ heterozygotes only ~1/3 of the CRCs harbour mutations in the A8 tract, ^2,3,11,12^ and co-occurrence of the two somatic *APC* hits (c.3924_3925insA and LOH of the wildtype allele) is identified in approximately 10% of the CRCs,^3,11^ suggesting that many of these tumours are not related to the germline *APC* I1307K variant. This observation is entirely consistent with the modest OR for association with CRC of germline I1307K: where the magnitude of association and accordant attributable fraction are modest, absence of the classical aetiological molecular hallmarks in a significant proportion of tumours in individuals with the germline aberration is well-described in the context of different molecular phenomena (such as LOH and mutational signatures) in tumour types of low ‘aetiologic index’.^13^ Nevertheless, along with population cohorts, such as UKBiobank, typically collecting constitutional DNA only, this factor would render investigation of tumours in non-AJ white I1307K variant carriers even more logistically implausible.

Whilst analysis of a prospectively ascertained cohort obviates the potential for inflation from population structure (ancestry bias between cases and controls), the low frequency of both I1307K and of CRC in any broad prospective cohort study renders prohibitive the size of dataset required to provide a definitive answer to this question. UKBiobank is noteworthy for having restricted recruitment to individuals aged 40-69 in order to enhance rates of accrual of cases of late-onset diseases; this cohort has now attained 14-18 years of maturation and the frequency of CRC is only 2%. For cohorts of young age of recruitment, for example AllofUs, accrual of sufficient cases of CRC will require decades of follow-up even despite a larger overall cohort size. Assuming equivalent maturity, accrual and age structure to UK Biobank (i.e. a frequency of CRC of 2%), a cohort three times the size of UKBiobank (1,495,550 individuals) would be required to provide 80% power for demonstration at 95% significance of an odds ratio of 1.7 (assuming a baseline frequency of I1307K of 0.08%).

Early studies performed in AJ, and non-AJ Jews identified a common haplotype for carriers with I1307K, regardless of their ethnic origin, suggesting a common origin.^14,15^ In line with this, Niell et al. identified a common progenitor haplotype in I1307K individuals of Ashkenazi, Sephardi and Arab descent ascertained in Israel.^16^ It is highly plausible that the founder haplotype expanded to other communities that came in contact with AJs or other Jewish populations that carried I1307K, although to our knowledge, no haplotype analysis in non-Jewish individuals living outside Israel has been reported.

Clinical guidance regarding *APC* I1307K is currently rather contradictory. NCCN guidelines (v.3.2024) recommend “high-quality colonoscopy screening” every 5 years (beginning at age 40, or 10 years before the youngest age at CRC diagnosis in a first-degree relative when there is family history of CRC), for all *APC* I1307K heterozygotes regardless of ancestry. By contrast, the UK Cancer Genetics Group (UKCGG), a constituent group of the British Society of Genomic Medicine, recommended that the I1307K variant is not reported as part of NHS-funded diagnostic *APC* testing, regardless of the individual’s ancestry (based on evaluation in the context of health priorities, opportunity cost and equity of access within the UK NHS). The rationale for this position argued on the basis of the modest magnitude of CRC risk conferred by *APC* I1307K.^17^ By contrast, InSiGHT recommend that, enhanced screening for CRC in *APC* I1307K heterozygotes should be restricted to individuals of AJ ancestry (as per enhanced screening offered to individuals with a first-degree relative affected with CRC) but that there should be no enhanced screening for non-AJ I1307K individuals because the risk association has not been proven for non-AJ ethnicities.

Overall, it would appear most likely that there is a single I1307K haplotype that confers modest susceptibility to CRC (and possibly other cancers) but it’s frequency outside the AJ population is too low to demonstrate statistically significant association with cancers within a single cohort. Our data demonstrate that adequately powered, unbiased case-control analysis of I1307K to assess association with CRC in the non-AJ white population would require a cohort of a size and maturity not plausibly feasible for decades. It is thus rational that (i) actionability of I1307K (i.e., incorporation for CRC risk stratification and concomitant alteration in clinical management) should be determined on the basis of the (modest) associated risk (ii) if I1307K is deemed clinically actionable, strategies for testing I1307K may be informed by ancestry (on account of differential detection rates). It is not however immediately rational that the management in those with I1307K should be informed by their ancestry.

## CONFLICT OF INTEREST

None

## FUNDING

SA and CFR were supported by CG-MAVE, Cancer Research UK Programme Award [EDDPGM-Nov22/100004]. LV’s research activity is funded by the Spanish Ministry of Science and Innovation (Agencia Estatal de Investigación), co-funded by FEDER funds-a way to build Europe- [PID2020-112595RB-I00, AEI/10.13039/501100011033], Instituto de Salud Carlos III [CIBERONC CB16/12/00234, PMP22/00064], and Government of Catalonia [AGAUR 2021_SGR_01112, CERCA Program for institutional support].

## ACKNOWLEDGEMENTS

This research has been conducted using the UK Biobank Resource under Application Number 76689 (https://www.ukbiobank.ac.uk/). We thank Gabriel Capellá for critical reading of the manuscript.

## AUTHOR CONTRIBUTIONS

Conceptualization: C.T., L.V.; Data curation: S.A.; Formal analysis: S.A., C.F.R., A.L., C.T., L.V.; Investigation: S.A., C.F.R., C.T., L.V.; Funding acquisition: C.T., L.V; Writing-original draft: C.T., L.V; Writing-review & editing: all authors

## ETHICS STATEMENT

UK Biobank has approval from the National Health Services National Research Ethics Service (16/NW/0274) and the North West Multi-Centre Research Ethics Committee as a Research Tissue Bank approval (11/NW/0382). Each participant provided written informed consent. Permission to access and analyze UK Biobank data (Project Application Number 76689) was approved by the UK Biobank according to their established access procedures.

## DATA AVAILABILITY STATEMENT

All data used in the analyses are available from UK Biobank upon request (https://www.ukbiobank.ac.uk). Data and methods relevant to the study are included in the article or uploaded as supplementary information.

## Notes

### Competing Interest Statement

The authors have declared no competing interest.

## REFERENCES

1. Valle L, Katz LH, Latchford A, et al: Position statement of the International Society for Gastrointestinal Hereditary Tumours (InSiGHT) on. J Med Genet 60:1035–1043, 2023

2. Laken SJ, Petersen GM, Gruber SB, et al: Familial colorectal cancer in Ashkenazim due to a hypermutable tract in APC. Nat Genet 17:79–83, 1997

3. Gryfe R, Di Nicola N, Gallinger S, et al: Somatic instability of the APC I1307K allele in colorectal neoplasia. Cancer Res 58:4040–3, 1998

4. Azzopardi D, Dallosso AR, Eliason K, et al: Multiple rare nonsynonymous variants in the adenomatous polyposis coli gene predispose to colorectal adenomas. Cancer Res 68:358–63, 2008

5. Privé F, Aschard H, Carmi S, et al: Portability of 245 polygenic scores when derived from the UK Biobank and applied to 9 ancestry groups from the same cohort. Am J Hum Genet 109:12–23, 2022

6. Bycroft C, Freeman C, Petkova D, et al: The UK Biobank resource with deep phenotyping and genomic data. Nature 562:203–209, 2018

7. Manichaikul A, Mychaleckyj JC, Rich SS, et al: Robust relationship inference in genome-wide association studies. Bioinformatics 26:2867–73, 2010

8. Behar DM, Metspalu M, Baran Y, et al: No evidence from genome-wide data of a Khazar origin for the Ashkenazi Jews. Hum Biol 85:859–900, 2013

9. Van Hout CV, Tachmazidou I, Backman JD, et al: Exome sequencing and characterization of 49,960 individuals in the UK Biobank. Nature 586:749–756, 2020

10. Forkosh E, Bergel M, Hatchell KE, et al: Ashkenazi Jewish and Other White APC I1307K Carriers Are at Higher Risk for Multiple Cancers. Cancers (Basel) 14, 2022

11. Zauber NP, Sabbath-Solitare M, Marotta S, et al: Clinical and genetic findings in an Ashkenazi Jewish population with colorectal neoplasms. Cancer 104:719–29, 2005

12. Zauber P, Bishop T, Taylor C, et al: Colorectal tumors from APC*I1307K carriers principally harbor somatic APC mutations outside the A8 tract. PLoS One 9:e84498, 2014

13. Hughley R, Karlic R, Joshi H, et al: Etiologic Index - A Case-Only Measure of BRCA1/2-Associated Cancer Risk. N Engl J Med 383:286–288, 2020

14. Shtoyerman-Chen R, Friedman E, Figer A, et al: The I1307K APC polymorphism: prevalence in non-Ashkenazi Jews and evidence for a founder effect. Genet Test 5:141–6, 2001

15. Patael Y, Figer A, Gershoni-Baruch R, et al: Common origin of the I1307K APC polymorphism in Ashkenazi and non-Ashkenazi Jews. Eur J Hum Genet 7:555–9, 1999

16. Niell BL, Long JC, Rennert G, et al: Genetic anthropology of the colorectal cancer-susceptibility allele APC I1307K: evidence of genetic drift within the Ashkenazim. Am J Hum Genet 73:1250–60, 2003

17. McVeigh TP, Lalloo F, Frayling IM, et al: Challenges in developing and implementing international best practice guidance for intermediate-risk variants in cancer susceptibility genes: APC c.3920T>A p.(Ile1307Lys) as an exemplar. J Med Genet 61:810–812, 2024

