## Supplementary Table 1, Supplementary Table 2, and Supplementary Figure 1 for "*APC* I1307K and clinical management: insights from UK Biobank association analysis of colorectal and other cancer risks in Ashkenazi and non-Ashkenazi whites"

### SUPPLEMENTARY MATERIAL

**Supplementary Table 1:** ICD9, ICD10 codes and self-reported diagnosis terms used to identify individuals diagnosed with each cancer phenotype.

| | ICD9 Codes | ICD10 Codes | Self-reported search term |
|---|---|---|---|
| ***Colorectal Cancer*** | 153.0, 153.1, 153.2, 153.3, 153.4, 153.5, 153.6, 153.7, 153.8, 153.9, 154.0, 154.1 | C18.0, C18.1, C18.2, C18.3, C18.4, C18.5, C18.6, C18.7, C18.8, C18.9, C19, C20 | “colo”, “rect”, “sigmoid”, “bowel” |
| ***Breast Cancer*** | 174.0, 174.1, 174.2, 174.3, 174.4, 174.5, 174.6, 174.8, 174.9, 175.0, 175.9 | C50.0, C50.1, C50.2, C50.3, C50.4, C50.5, C50.6, C50.8, C50.9 | “breast” |
| ***Prostate Cancer*** | 185 | C61 | “prostate” |
| ***Pancreatic Cancer*** | 157.0, 157.1, 157.2, 157.3, 157.4, 157.8, 157.9 | C25.0, C25.1, C25.2, C25.3, C25.4, C25.7, C25.8, C25.9 | “pancrea” |
| ***All Cancers*** | 140-165, 170-176, 179-209, 230-239 | C00-C97, except C44 (non-melanoma skin cancer) | All indications except “non-melanoma skin cancer” |

**Supplementary Table 2:** *APC* I1307K frequency in breast, prostate, pancreatic, and all cancer patients and patients without these phenotypes in the UK Biobank, and associated ORs for AJ and non-AJ individuals.

| Ancestry | Cancer patients (I1307K/total) | Individuals w/o the corresponding cancer type (I1307K/total) | OR (95% CI) | Fisher's exact p-value |
|---|---|---|---|---|
| | ***Colorectal cancer*** | | | |
| ***AJ*** | 2/39 (5.1%) | 191/2687 (7.1%) | 0.71 (0.17 – 2.95) | 1 |
| ***Non-AJ White*** | 7/8688 (0.08%) | 329/428250 (0.08%) | 1.05 (0.50 – 2.22) | 0.84 |
| ***Non-AJ Black*** | 0/85 (0.0%) | 2/7158 (0.02%) | *NA* | *NA* |
| ***Non-AJ Asian*** | 0/73 (0.0%) | 13/8980 (0.1%) | *NA* | *NA* |
| ***Non-AJ Chinese*** | 0/15 (0.0%) | 0/1432 (0.0%) | *NA* | *NA* |
| ***Non-AJ Mixed*** | 0/44 (0.0%) | 9/2653 (0.3%) | *NA* | *NA* |
| | ***All cancers*** | | | |
| ***AJ*** | 45/538 (8.4%) | 148/2188 (6.76%) | 1.26 (0.89 – 1.78) | 0.19 |
| ***Non-AJ White*** | 62/87705 (0.07%) | 274/349233 (0.08%) | 0.90 (0.68 – 1.19) | 0.50 |
| ***Non-AJ Black*** | 0/894 (0.0%) | 2/6349 (0.03%) | *NA* | *NA* |
| ***Non-AJ Asian*** | 1/850 (0.1%) | 12/8203 (0.1%) | 0.80 (0.10 – 6.19) | 1 |
| ***Non-AJ Chinese*** | 0/156 (0.0%) | 0/1291 (0.0%) | *NA* | *NA* |
| ***Non-AJ Mixed*** | 0/375 (0.0%) | 9/2322 (0.4%) | *NA* | *NA* |
| | ***Breast cancer*** | | | |
| ***AJ*** | 11/131 (8.4%) | 182/2595 (7.0%) | 1.22 (0.64 – 2.29) | 0.49 |
| ***Non-AJ White*** | 14/18683 (0.07%) | 322/418255 (0.08%) | 0.97 (0.57 – 1.66) | 1 |
| ***Non-AJ Black*** | 0/167 (0.0%) | 2/7076 (0.03%) | *NA* | *NA* |
| ***Non-AJ Asian*** | 0/232 (0.0%) | 13/8821 (0.1%) | *NA* | *NA* |
| ***Non-AJ Chinese*** | 0/48 (0.0%) | 0/1399 (0.0%) | *NA* | *NA* |
| ***Non-AJ Mixed*** | 0/102 (0.0%) | 9/2595 (0.4%) | *NA* | *NA* |
| | ***Prostate cancer*** | | | |
| ***AJ*** | 7/85 (8.2%) | 186/2641 (7.0%) | 1.18 (0.54 – 2.60) | 0.67 |
| ***Non-AJ White*** | 7/14159 (0.05%) | 329/422779 (0.08%) | 0.64 (0.30 – 1.34) | 0.28 |
| ***Non-AJ Black*** | 0/312 (0.0%) | 2/6931 (0.03%) | *NA* | *NA* |
| ***Non-AJ Asian*** | 1/137 (0.7%) | 12/8916 (0.1%) | 5.46 (0.70 – 42.3) | 0.18 |
| ***Non-AJ Chinese*** | 0/11 (0.0%) | 0/1436 (0.0%) | *NA* | *NA* |
| ***Non-AJ Mixed*** | 0/46 (0.0%) | 9/2651 (0.3%) | *NA* | *NA* |
| | ***Pancreatic cancer*** | | | |
| ***AJ*** | 1/9 (11.1%) | 192/2717 (7.1%) | 1.64 (0.20 – 13.21) | 0.48 |
| ***Non-AJ White*** | 0/1458 (0.0%) | 336/434808 (0.08%) | *NA* | *NA* |
| ***Non-AJ Black*** | 0/19 (0.0%) | 2/7224 (0.03%) | *NA* | *NA* |
| ***Non-AJ Asian*** | 0/15 (0.0%) | 13/9038 (0.1%) | *NA* | *NA* |
| ***Non-AJ Chinese*** | 0/3 (0.0%) | 0/1444 (0.0%) | *NA* | *NA* |
| ***Non-AJ Mixed*** | 0/8 (0.0%) | 9/2689 (0.3%) | *NA* | *NA* |

**Supplementary Figure 1:** Ethnicity breakdown of UK Biobank individuals included in analysis. Breakdown is by top-level ethnicity, and by PC-assessed AJ ancestry status.

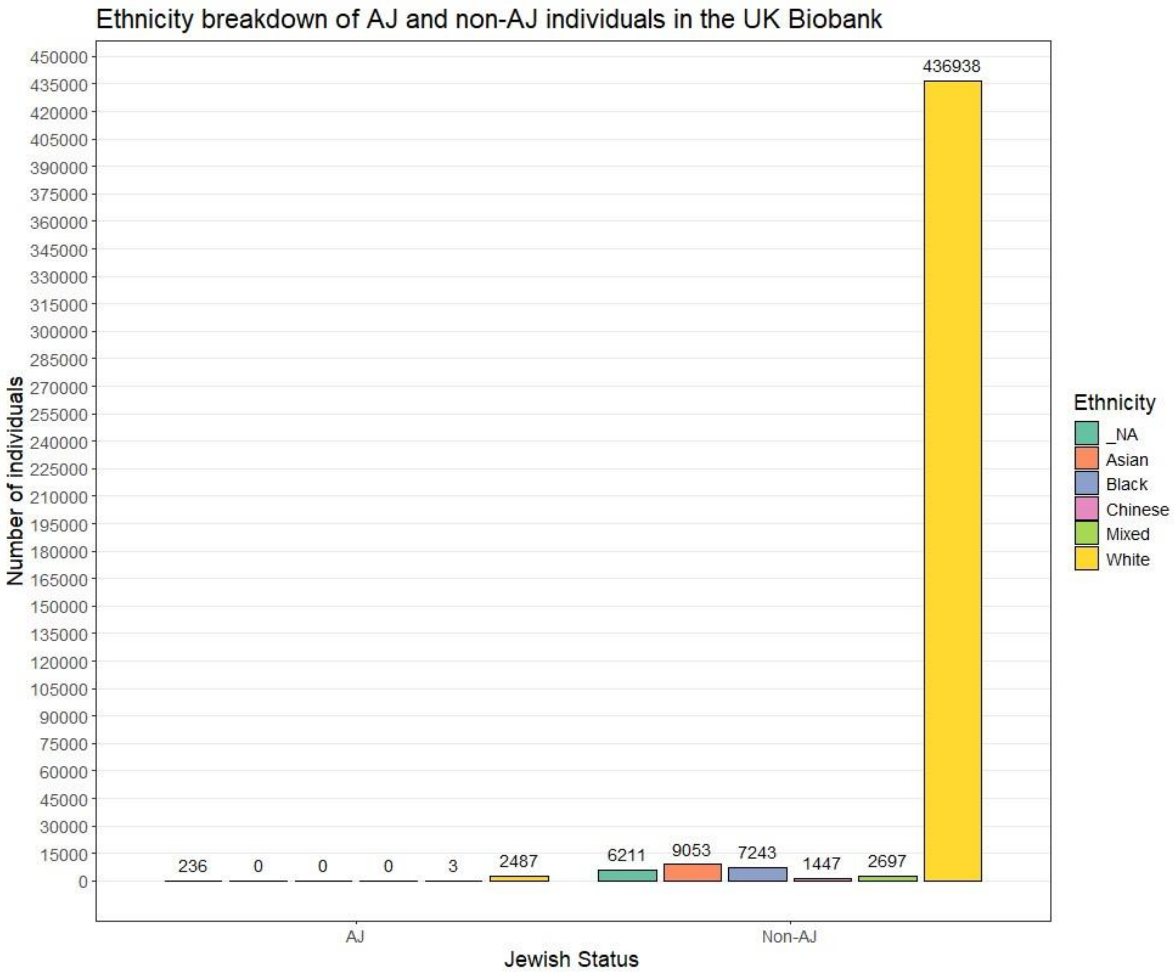
